# Years of life lost due to the psychosocial consequences of COVID19 mitigation strategies based on Swiss data

**DOI:** 10.1101/2020.04.17.20069716

**Authors:** Dominik A. Moser, Jennifer Glaus, Sophia Frangou, Daniel S. Schechter

## Abstract

**Background:** The pandemic caused by COVID-19 has forced governments to implement strict social mitigation strategies to reduce the morbidity and mortality from acute infections. These strategies however carry a significant risk for mental health which can lead to increased short-term and long-term mortality and is currently not included in modelling the impact of the pandemic.

**Methods:** We used years of life lost (YLL) as the main outcome measure as applied to Switzerland as an exemplar. We focused on suicide, depression, alcohol use disorder, childhood trauma due to domestic violence, changes in marital status and social isolation as these are known to increase YLL in the context of imposed restriction in social contact and freedom of movement. We stipulated a minimum duration of mitigation of 3 months based on current public health plans.

**Results:** The study projects that the average person would suffer 0.205 YLL due to psychosocial consequence of COVID-19 mitigation measures. However, this loss would be entirely borne by 2.1% of the population, who will suffer an average 9.79 YLL.

**Conclusions:** The results presented here are likely to underestimate the true impact of the mitigation strategies on YLL. However, they highlight the need for public health models to expand their scope in order to provide better estimates of the risks and benefits of mitigation.

## Introduction

Coronavirus disease 2019 (COVID-19) has led to the first truly global pandemic. At the time of writing this paper, there were over two million reported cases worldwide and more than 130,000 deaths attributed to COVID19 acute infection (1). Based on models of its spread, and potential for morbidity and mortality, most governments worldwide have adopted mitigation strategies that essentially limit social contacts (2, 3). The goal of these measures is to “flatten the curve” of acute presentations so as to prevent widespread morbidity and the break-down of health care systems. Variants of these social mitigation strategies range from “social distancing”, at-home-confinement-referred to as “self-isolation”, to selective “quarantine”, and to population “lockdown” that includes restriction of movement outdoors and closure of schools and all non-essential services and businesses.

None of the existing models have factored the possible adverse mental health effects of mitigation at a population level. These adverse effects can be intuitively anticipated but have never been rigorously modelled (4). Negative mental health outcomes can be attributed to the emotional and physiological effects of the risk posed by the virus and by reduced physical activity, social interaction and human physical contact (5–7). Studies on prior pandemics, such as the Severe Acute Respiratory Syndrome (SARS)found that the length of quarantine was an important predictor of post-traumatic stress disorder (PTSD), depression and anxiety with a cumulative prevalence exceeding 30% of the population (8, 9). Psychosocial stressors within families and loneliness for those living alone are also likely to spike in confinement and have adverse effects on mental and physical health (10–13). Available data suggest that stress associated with population-wide disasters increases the level of violence, including domestic violence and child abuse (14, 15). These are recognised risk factors for mental health and substance abuse problems (16) as well as suicide (17).

The anticipated impact of the COVID19 pandemic on mental health is expected to be significant but has not been considered in formulation current public policies. To address this gap, the present study makes a rapid model-projection concerning the years of life lost (YLL) if restrictive social mitigation measures are implemented for a period of 3 months. This duration was chosen as it aligns with the expected duration of social mitigation in many countries. We use data from Switzerland as an example. The model focuses on what we consider to be the major contributors to YLL affecting the majority of the population, namely: suicide, emergence or increase in psychopathology, childhood physical abuse and continued restriction of movement and at home confinement.

To be clear, this model focuses on changes to psychosocial risk factors. The COVID-19 crisis may also have other adverse consequences that may impact on longevity such as economic adversity, changes to activities of daily living such as eating, sleeping, smoking and ordinary alcohol consumption or decrease in medical provision to those who have health problems unrelated to COVID-19. Such additional factors are however beyond the scope of the present study. A more precise estimation of the mental health impact of the pandemic will be possible as relevant data become available.

### Methods

#### Model

We conducted a literature review focusing on studies reporting on YLL in connection to situations conceptually similar to the current pandemic. These included data from studies on confinement in different contexts and from previous disasters including pandemics. We focused on studies from developed countries, primarily Switzerland, and when not available, from Europe followed by the United States, based on the United Nations Development Programme-Country Classification System. Switzerland has a population of 8.57 million (18) and introduced an “extraordinary situation” on March 16 2020. All boarders were closed to travel, all schools, markets, restaurants, non-essential shops, bars and entertainment and leisure facilities were closed, and all public and private events and gatherings were prohibited (19). Several regions had already taken a number of these measures in the preceding days and weeks. The Federal Council called on members of the public to avoid all unnecessary contact, maintain physical distance from others and stay whenever possible at home. According to government announcements these measures are expected to continue until at least April 27 2020 while a number of measures aimed at “social distancing”, including prohibition of gatherings, are expected to last until at least June 8 (20).

The following risk factors were considered based on their importance and data availability: Suicidality, depression, alcohol use disorder, childhood trauma due to domestic violence, changes in marital status, and social isolation. The projection of YLL for each of these factors is further described below. Data concerning the incidence of the risk factors as well as their impact on YLL were then applied to a model that assumes population-wide severe social mitigation policies (stay-at-home and restriction of outdoors movement) for a duration of 3 months. As a general rule, the present model erred on the conservative side when choosing YLL. For purposes of illustration only, we also present projected YLL for countries other than Switzerland based on their population size (21) assuming similar prevalence of risk factors.

The model involved a six step process.

For each factor:

1. ***Estimation of baseline risk of outcome i (BRi) based on the literature***
2. ***Estimation of YLL per incident of outcome i (YLLi) from the literature***
3. ***Estimation of increased risk factor during the pandemic for outome i (PRi), where possible based on literature***
4. ***Estimation of the increased incident cases relating to the pandemic outcome i (PICi)*** PIC_i_=(PRi-1)*BRi*0.25 Where PR_i_ is the estimate of the increased risk of outcome *i* relating to the pandemic, D is the duration of the social mitigation measures, which is fixed 0.25 years (3 months)_i_
5. ***Estimation of YLL for incidencei due to the pandemic (PYYLi)*** PYLL_i_= PICi *YLLi
6. ***Calculation of summary statistics*** PICs is the sum of all PICi; PYLLs is the sum of all PYLLi Average YLL per impacted Person: PICs / PYLLs Percentage of Persons impacted: PICs/100* Population of Switzerland (8.57 million) Average PYLL per person of the general population: PYLLs/ Population of Switzerland (8.57 million)

To align with current models that focus on acute mortality, we focus on the 3-month period which represents an underestimate of the overall impact of the pandemic.

## Results

A summary of the results is provided in Table 1 and details of the estimation of the increased YLL linked to the pandemic are presented below.

**Table 1:** Projection of lost years of life for the population of Switzerland due to demographics and mental health changes related to a mass confinement of 3 months.

| Cause | Estimated Persons Affected with reduced longevity (% of the overall population) | Years of Life Lost per Person | Years of Life Lost for an isolation 3 months period | Years of life lost per person across the entire population (8.57 million) |
| --- | --- | --- | --- | --- |
| Suicidality | 1'523 (0.02%) | 34.3 | 52'239 | 0.006 |
| Divorces (spouses) | 20'842 (0.24%) | 3.5 | 72'947 | 0.008 |
| Divorces (affected minors) | 7'694 (0.09%) | 4 | 30'776 | 0.004 |
| Family Violence (affected minors) | 5'567 (0.06%) | 2.37 | 13'194 | 0.002 |
| Depression | 94'613 (1.10%) | 6.82 | 645'260 | 0.075 |
| Alcohol use Disorder | 51'000 (0.60%) | 17.67 | 901'170 | 0.105 |
| Diminished Social contacts | 3'484 (0.04%) | 12.03 | 41'912 | 0.005 |
| Total | 179'520 (2.09%) | 9.79 | 1'757'498 | 0.205 |

**Table 2.** indicates how many Years of life lost selected other countries would be projected to have, were their disorder and social representation were the same as Switzerland’s (it is simply a multiplication of the years of life lost by the size of the population). Population numbers for countries other than Switzerland were sourced from Wikipedia (21).

| Country | Population in millions | Years of Life Lost for an isolation 3 months period in millions |
| --- | --- | --- |
| Switzerland | 8.57 | 1.76 |
| Germany | 83.15 | 17.05 |
| France | 67.08 | 13.75 |
| Spain | 47.10 | 9.65 |
| United Kingdom | 66.44 | 13.62 |
| Canada | 37.99 | 7.79 |
| United States | 329.61 | 67.58 |
| Japan | 125.95 | 25.82 |
| China | 1402.16 | 287.47 |

### Suicidality

BRi: In 2017, Switzerland counted 1043 suicides excluding assisted suicide or euthanasia. Non euthanasia-related suicide was the cause of death in 16 out of 1000 deaths (22).

YLLi: US data from 2016, showed 1.542 million years were lost due to suicide (23) across 44965 suicides (24) leading to 34.3 YLL per death (1.542 million/44965).

PRi: We extrapolated on the relationship between confinement and suicidality from data from the penitentiary system. Community cell-confined prisoners already have an increased suicide rate by factors between 3.5 and 21 compared to the general population (25, 26). However, the risk of suicide for prisoners in single cells is further increased approximately 9 to 15 times (27). Extrapolating from these data, we assume that confinement in the household increases their likelihood of suicide by a factor of 3 in multi-person households and a factor of 27 in single-person households (3*9). We assume this increase is stable for the entire 3-month duration. In Switzerland, 16% of the population live in single-households (28). These calculations result in a population-wide PRi of 6.84 (0.16*27 + 0.84*3).

PICi: 5.84* 1043*0.25= 1523 additional suicides.

PYLLi: = 1523*34.3 = 52239.

### Depression

General: As mood and anxiety disorders are comorbid, we used data on depression in our model as it is likely to capture much of the distress related psychopathology; additionally, depression has the most convincing link to YLL which is our outcome of interest (29).

BRi: We used population prevalence data for depression as they capture both incident and pre-existing conditions(30, 31). Accordingly, we estimated that the pre-pandemic risk of depression for the Swiss population in a 3-month period is 3.45% (i.e. 8.57 million*0.0345= 295’665) with 64.7% of affected individuals being women. Because BRi is already adjusted for the 3-month period, no further correction for PICi was undertaken.

YLLi: Using data from a prior study on depression (32), we assumed loss of 7.91 years of life for men and 6.22 for women. Given the male-female ratio for depression (64.7% women) this results in YLLi = 6.82.

PRi: Three years after the SARS epidemic, the proportion of persons with symptoms related to higher stress was still increased by a factor of 3.47 among those who had been in quarantine, thus demonstrating the long-term implications of the phenomenon (33). A study of an Australian population quarantined due to equine flu suggested a similar 3-fold increase in depression (34). We used the latter estimate as it was the most conservative. We also factored in that -given therapy- 84% of individuals with depression are likely to remit within 3 years (31). To be conservative and to capture cases most likely associated with mortality, we adjusted the model accordingly leading to a PRi of 1.32.

PICi: 295665*0.32=94613.

PYLLi: 94613*6.82= 645’2260 YLL.

### Alcohol use disorder

General: Distress under any circumstances is a known risk factor for alcohol abuse disorder (AUD). There is abundant evidence of increased alcohol consumption during the current pandemic (35) including a report of approximately one third increase in alcohol sales in Germany (36). Although the use of other substances seems to be increased as well, our model focuses on AUD as it is the most prevalent substance abuse disorder and the major contributor to mortality worldwide (37). Substance use disorders are significantly associated with increased mortality due to increased accidents, impulse-dyscontrol leading to violence and suicide, as well as increased physical morbidity (i.e. cardiovascular, gastro-intestinal, hepatic, and other somatic conditions) (38-40).

BRi: In Switzerland, 16.1 of men and 3.2% of women were estimated to suffer from AUD, meaning that 83.5% of cases are men and 16.5% are women (37).

YLLi: A cross-national Scandinavian study indicated that life expectancy among inpatients with AUD was reduced by 24-28 years compared to the general population (40). When only deaths from natural causes are considered, then life expectancy is reduced by 18.1 years in men and 16.5 years in women (39). To be conservative, and to account for the preponderance of men with AUD we assumed a reduction of life expectancy by 18 years in men (83.5% of cases) and 16 years in women (16.5% of cases) resulting in a total YLLi of 17.67 years.

PRi: We assume a population-level increase in AUD of 0.15% per month, with the first month leading to a higher incidence (0.3%). Therefore, countermeasures lasting 3 months would increase incidence by approximately 0.6%.

PICi: 0.6% of the population (8.57 million) = 51’000.

PYLLi: 51000*17.67 = 901’170.

### Marital Status

General: Recent media reports indicate that divorce rates have increased since the instigation of COVID-19 mitigation policies (11, 41) In individual cases, divorces and separations can be beneficial to individual health and stress levels (for example in situations of abuse). Overall, however, even after taking into account risk factors that contribute to divorce and separation (i.e. financial stressors, mental and physical illness and substance abuse), divorce and separation given the experience of relational and economic stress, loss, and greater likelihood for social isolation, have been shown to have a negative impact on longevity (10). Causes for this may also include detrimental habits that individuals may adopt to cope with the stress, loss, and isolation (such as increased smoking (42)). Additionally having parents who divorce during childhood has been estimated to increase mortality by 44% and reduce life expectancy by an average of 4 years (43).

BRi: In 2018, there were 16542 divorces in Switzerland leading to a BRi=33084. Additionally 12212 minors were affected by the breakdown of marital relationships (44).

YLLi: A German study estimated that YLL atteibutable to divorce range between 3-8 years for women and 4-9 years for men (45). For this projection, 3.5 YLLi were modeled per couple (4 years for men, 3 years for women) and 4 years for each affected minor (43).

PRi: We based our calculation on the increase in divorce rate for the year following the Hurricane Hugo disaster (46) (wherefore factor D is omitted in the calculation of PICi). PRi was modelled as 1.63.

PICi: for adults: 33084 *0.63 = 20842; for affected minors: 12212*0.63 = 7694.

PYLLi: for adults: 20842*3.5= 72947; for affected minors: 7694*4=30776.

### Childhood trauma due to domestic violence

General: Although family violence is commonly targeted towards both women and children, we focus specifically on the effects on children as specific impact on women was hard to quantify.

BRi: Even when not directly being the victim themselves, children being witnesses to violence can be an adverse childhood event.

In 2013, 9381 victims of domestic violence registered by the Swiss police. However, a survey indicated that that this latter number of victims would only represent 22% of the actual number, which would increase the number to 42641 victims per year (9381/22*100). 64.5% of domestic violence referred to violent interactions between either parents and their children or current romantic partners (47) (42641*0.645=27’503). Of Swiss multi-person-households 46% include children (28) and on average there are 1.76 children living in each of these households (48). BRi is therefore assumed to be 22’266 (27503*0.46*1.76).

YLLi: Experiencing 3 or more adverse childhood events (ACEs) is associated with 9.5 years of reduced expected quality longevity (49). Among adults, 25% report having experienced multiple averse childhood events (50). Because of this we conservatively project that only about every fourth of these events will lead to the full loss of 9.5 years; we therefore adjusted the YLLi to 2.37 years.

PRi: According to the World Health Organization (WHO), there has been a threefold increase in family violence since the start of the pandemic (51). However, additional events are not likely to be normally distributed across victims (47) and the measures to which these numbers refer may have been stricter than the one in Switzerland; accordingly we adopted a conservative PRi = 2.

PICi: 22266*1*0.25=5567.

PYLLi: 5567*2.37=13194.

### Social isolation and reduced social connectedness

General: No studies were found that indicated the cost of social isolation or reduced social connectivity in YLL in a way directly adaptable to the present study. Moreover, the entire population is somewhere on a spectrum from socially hyperconnected to socially isolated. However, studies concerning risk ratios do exist.

BRi: The entire Swiss population of 8.57 million is on a spectrum from socially connected to socially isolated, depending on their personal circumstances.

YLLi: Based on the most recent data, Switzerland counted 67008 deaths in 2018 that were distributed across age groups as follows: category 1 (ages: 0-19): 0.8%; cat 2 (20-39): 1.2%; cat3 (40-64): 11.1%; cat 4 (65-79): 25.0%; cat5 (80+): 61.9% (22). There are no data informing on the baseline number of deaths that can be attributed to social disconnection or loneliness. In response, we averaged the life-expectancy of men and women and calculated YLLi according to the following steps: 1) life expectancy by category was taken from the Federal Statistics office of Switzerland which gives remaining life expectancy at birth, 30, 50, 65 and 80 years of age. (52). 2) We then conservatively adjusted YLL for each age category as a very rough approximation. This approximation is -if anything- aimed at underestimating the remaining life expectancy (cat 1: 73.45, cat 2: 54.2, cat 3: 34.8, cat 4: 15.5, cat 5: 4.95). 3) Overall, remaining life expectancy was then multiplied with the percentage of deaths in each age group, leading to a cumulative YLLi =12.03 for each additional death.

PRi: Having more social connections has been associated with lower death rate with an odds ratio of 1.5 (53). Conversely, a comprehensive meta-analysis indicated that social isolation is associated with an increase of all-cause mortality by a factor of 1.29 (54). Similarly, a large-scale study estimated that social isolation increased the hazard risk by a factor of 1.26 after adjusting for multiple potential confounders including anxiety, depression and lower socio-economic status (55). In our model, we use the most conservative estimate of 1.26. In a phase of confinement, we assume that a majority of 75% of the population experiences either reduced social connectedness and/or increased social isolation. This would lead to a PRi of 1.208.

PICi: Based on the 2018 data on the number of deaths in Switzerland (n=67008) (22), PICi was estimated as 67008*0.208*0.25=3484.

PYLLi: 3484*12.03=41’912.

### Summary Statistics

The findings presented above are summarized in Table 1. The contribution of the different risk factors to PYLL in descending order was: Alcohol use disorder: 901’170, Depression: 645’260, Divorce: 103’723, Suicide: 52’239, Reduction of Social contact: 41’912, Averse childhood events due to domestic violence: 13’194.

The sum of all PYLLi was 1’757’498, this represents 0.205 PYLL per person in Switzerland (1757498/8.57 million) (Table 1). In other words, we project a loss of 10 weeks and 5 days due to COVID-19 related mitigation strategies if YLL is equally distributed in the entire Swiss population. The sum of all PICi was 179’520 which represents 2.1% of the Swiss population (179520/8.57 million). Assuming that this subpopulation will be most impacted, the average PYLL was estimated to be 9.79 (1757498/179’520=9.79).

## Discussion

The current study focused on years of life lost due to the social mitigation strategies implemented in response to the COVID19 pandemic, with a primary focus on the consequences of at home confinement and restriction to freedom of movement.

The literature suggests that increased duration of confinement is associated with worse outcomes for psychological health of those confined (4). While some of the stress related problems ensuing from confinement may remit, an important portion of this damage may prove to be hard or impossible to reverse and the affected individuals may experience on going suffering. Our projection suggests that the Swiss population will incur a substantial increase in mortality as a consequence of confinement related psychosocial stress, which should be considered in forming public health responses to the pandemic. It is important that policy makers factor mental health when conduction cost-benefit analyses of mitigation strategies.

The present study hopes to have achieved three aims: 1) to provide information that helps authorities to consider whether and, if so, how to enact these countermeasures and what resources to employ for mitigation of their adverse consequences. 2) specifically indicate the importance of support for mental health care workers in order to allow them to be maximally efficient in the face of confinement measures 3) to make the case for more comprehensive modelling of the effect of pandemic-responses beyond the immediate risk attributed to acute infection.

Limitations: As we demonstrate here, the evidence base for building such comprehensive models is limited and accordingly we had to make several assumptions. In this sense, our model projection is obviously constrained by the limitations of the available literature which, itself, involves a number of unknowns. Given the time constraints, the uncertainty in those assumptions is increased; the authors however judge the urgency of such projections to be very high at the moment. Moreover, in this respect our model is not dissimilar to current projections of the spread and consequences of COVID19 which are being continually revised as more. Additionally, the present projection is not all-encompassing concerning potential effects of confinement: such as (prolonged) grief, elder abuse, increase of sedentary lifestyle or change of diet. The pandemic is also likely to have multiple additional consequences including distress due to job losses and financial hardship. The projection also does not model potentially positive changes in behaviour, for example reductions in car accidents and air pollution. Due to frequent co-occurrence of certain phenomena it is possible that a single individual may be affected by more than one of the factors presented. When possible, data were adjusted for age, sex and socioeconomics. However, for several factors, possibilities to do so were impeded by virtue of limitations of the current literature.

## Data Availability

All data is available within the manuscript itself

